# Home polysomnography in children with autism spectrum disorder: a prospective observational study

**DOI:** 10.1101/2025.09.19.25336196

**Authors:** Uchenna Ezedinma, Scott Burgess, Janet Greenhill, Jyoti Singh, Evan Jones, Andrew Ladhams, Gary Campell, Shauna Fjaagesund, Piotr Swierkowski, Alexandra Metse, Terri Downer, Florin Oprescu

## Abstract

This prospective observational study reports on the feasibility and adequacy of feasibility and adequacy of Level 2 polysomnography involving children with autism spectrum disorder during an interventional-randomised controlled trial.

Multiple level 2 polysomnographic studies were performed using Nox-A1 devices worn between October 2023 and September 2024. Study feasibility was determined by the child’s compliance and primary caregiver report, while signal quality (key channels present for at least 90% of sleep time) was used to define study adequacy. A cost analysis was also conducted.

Twenty children (6-12 years, 9.1+1.55 years: 16 males) with autism spectrum disorder (level 2) and reported sleep difficulties participated in the study.

Eighty (89%) of 90 polysomnographic studies were feasible. All infeasible studies, except one, were unrelated to the study. Seventy-four (93%) of the eighty studies resulted in adequate study quality. Most (n=6, 7%) inadequate studies were due to electroencephalogram signal artefact/absence. The participants did not have a sleep disorder requiring medical attention. The cost of a study was estimated at $AUD 258.

The study indicates the feasibility, adequacy and cost-effectiveness of level 2 polysomnography in evaluating sleep outcomes in children with autism spectrum disorder during an interventional randomised controlled trial. This preliminary study provides valuable insights into the field of paediatric sleep medicine. Repeat studies of this method using diverse and larger sample sizes are warranted.

## Introduction

Sleep difficulties are estimated to occur in 50–80% of children with autism spectrum disorder (ASD) compared to 11–37% in typically developing children (Rana et al., 2021). Frequently reported sleep difficulties co-occurring in ASD include bedtime resistance, difficulties falling asleep and staying asleep, sleep fragmentation, and early morning awakenings (Rana et al., 2021, Maurer et al., 2023)— these impact neural development, cognition, and behaviour (Feng et al., 2021). Therefore, research into potential interventions within the population is of priority (Mindell et al., 2009). These potential interventions are generally evaluated using several sleep assessment measures (Banaschewski et al., 2022). However, there is sparse evidence on the use of sleep studies within interventional research involving children with ASD (Hodge et al., 2012, Moore et al., 2017, O’Sullivan et al., 2023).

Sleep studies are valuable for assessing overlapping or complicated sleep difficulties (Douglas et al., 2017), especially in autistic individuals who may experience multiple and concurrent sleep difficulties. There are four levels of sleep studies, broadly classified into two categories: Polysomnography (levels 1 & 2) and Limited channel sleep studies (levels 3 & 4). Generally, polysomnography (PSG) provides a comprehensive sleep assessment from multiple sleep parameters, such as electroencephalography, electrooculography, electrocardiography, electromyography, anterior tibial muscle activity, arterial oxygen saturation, sound, respiratory thoraco-abdominal movements, airflow (mouth and nose thermistors) and body position. These allow for assessing sleep stages and accurately quantifying respiratory events (against time spent asleep) in diagnosing sleep difficulties (Douglas et al., 2017).

There are differences between level 1 PSG (L1PSG) and level 2 PSG (L2PSG). A L1PSG is a laboratory-based sleep study attended by trained sleep laboratory staff, while a L2PSG is an unattended PSG study (no medical staff present) and is performed outside of a laboratory, typically in the patient’s home. The presence of trained sleep laboratory staff, coupled with synchronised video monitoring, minimises artifacts in L1PSG, as the staff can address issues in real-time during the study, thereby ensuring high-quality signal acquisition compared to L2PSG (Portier et al., 2000). Also, in L1PSG, the controlled laboratory setting minimises external disturbances, thereby improving accurate data collection, while in L2PSG, external disturbances like noise or family interaction could potentially impact the data collection and quality. L1PSG is the “gold standard” against which L2PSG is assessed.

Notwithstanding, there are some advantages of L2PSG over L1PSG. The latter is labour-intensive and costly due to the need for trained sleep laboratory staff. For instance, in a cost comparison, L2PSG was demonstrated to be one-third of the current cost of L1PSG (Carpentier et al., 2014). More so, some studies suggest L1PSG induces an atypical or poorer sleep - a *first-night effect* (Aathira et al., 2017, Buckley et al., 2013). A recent multi-centre study reported that L1PSG was non-representative of usual sleep at home, especially for children with autism spectrum / neuromuscular disorder (Kevat et al., 2025). In a bid to minimise the *first-night effect*, some studies have used an adaptation night or two consecutive L1PSG (Miano et al., 2007, Newell et al., 2012) or a calming strategy involving multiple staff (Bessey et al., 2013). Paradoxically, these modifications to the L1PSG method would be more labour-intensive and costly.

On the contrary, L2PSG is not only less labour-intensive and costly, but a single L2PSG is feasible (Carpentier et al., 2014, Brockmann et al., 2013, Russo et al., 2021), without any *first-night effects* (Pedersen et al., 2023). A recent systematic review concluded that L2PSG is safe and technically feasible with relatively low failure rates in adults (Braun et al., 2024). In children, a majority of primary caregivers reported the child had typical or better sleep, and that L2PSG was easy or very easy (Russo et al., 2021). There is a higher preference for L2PSG over L1PSG (Kevat et al., 2025). This may be because children with neurodevelopmental conditions such as ASD are less likely to tolerate L1PSG, primarily due to poor tolerance to the nasal prongs and oronasal thermistor needed for measuring direct airflow for the assessment of sleep disordered breathing, such as obstructive sleep apnea (Lanzlinger et al., 2023).

There is a need to develop supportive practices and alternative measures to L1PG (Lanzlinger et al., 2023), if sleep studies are to be used in interventional research involving children with ASD. L2PSG may be considered given its advantages compared to L1PSG. Current L2PSG studies have involved typically developing children (Pedersen et al., 2023, Goodwin et al., 2001); and when L2PSG were investigated in children with ASD, studies used direct airflow monitoring (Kawai et al., 2022) or were set up in a laboratory (Russo et al., 2021, Maski et al., 2015) or included a desensitisation technique (Kawai et al., 2022). The direct airflow monitoring may have underpinned the hypothesised sensory sensitivities and intolerance (Gao et al., 2022) or the potential cost implication of two consecutive L2PSG (Jiao et al., 2022), resulting in its sparse use in interventional research.

Modification to L2PSG methods may optimise its use in interventional research involving children with ASD. For instance, the calibrated signal derived from bands worn around the chest and abdomen can be used to indirectly monitor airflow (Russo et al., 2021). Even with a 10% signal loss, such as the respiratory effort signals, L2PSG did not alter treatment advice compared to L1PSG (Campbell and Neill, 2011, Ghegan et al., 2006). Setting up in the patient’s home may increase the feasibility rate. In a comparative study between L1PSG and L2PSG, one-third of the patients reported difficulties transporting to the laboratory in the late afternoon for set up and back the next morning after sleeping in their home (Portier et al., 2000). It could be argued that setting up the patient in their own home may improve the feasibility of L2PSG as reported in a study by Withers et al. (2022).

Therefore, it could be hypothesised that L2PSG setup at home and using an indirect airflow measure is highly feasible and adequate during an interventional research involving children with ASD and sleep difficulties.

## Materials and method

### Settings

This study reports on polysomnographic data prospectively collected during an interventional randomised waitlist-controlled pilot study (Australian New Zealand Clinical Trials Registry (ANZCTR): 12623000757617). In phase 1 of the RCT, twenty participants were to receive a L2PSG before (T1) and after (T2) the intervention. In phase 2 of the RCT, ten participants were to receive a PSG study after the intervention (T3). In phase 3 of the RCT, the twenty participants were to receive a L2PSG study at one (T4) and four (T5) months post-intervention follow-up. The study was conducted between October 2023 and September 2024 at the participant’s home in Queensland, Australia. The protocol and informed consent forms in this observational study were prospectively received and approved by the University of Sunshine Coast Australia Ethics Committee in February 2023, with ethics number S221766.

### Participants

The participants were part of the interventional randomised controlled clinical trial. Briefly, they were recruited from the community via advertisements on relevant institutions’ notice boards, health consumer web pages and social media support groups that targeted primary caregivers of children with ASD. The relevant study inclusion criteria were children (6-12 years old) with a diagnosis of ASD (level 2) based on the Diagnostic and Statistical Manual of Mental Disorders, Fifth Edition, by a paediatrician and primary caregiver-reported sleep difficulties (>41 total scores on the Children Sleep Habit Questionnaire). Participant’s demographic data such as age, gender, and comorbidities were collected at baseline.

### Intervention

Briefly, the intervention was alpha rhythm (8-13Hz) guided repetitive transcranial magnetic stimulation (α-rTMS) sessions, commercially known as magnetic EEG-EKG resonant therapy (MeRT), during consecutive business days on a CF-B65 butterfly coil and Magpro R30 TMS stimulator (Magventure Inc, Denmark). The personalised stimulation frequency was processed using a proprietary algorithm that calculated and identified the participant’s alpha rhythm (8-13Hz range) within the posterior-occipital region of interest (ROI) consisting of P3, P4, Pz, O1, and O2 electrodes. The α-rTMS protocol was five seconds of stimulation (8.0-13.0Hz range), delivered first to the Pz site and then to the Fpz site, with 28-second intervals between 32 trains and at 40% and 50% output intensity for each site, respectively (Ezedinma et al., 2022). Each α-rTMS session was approximately 40 minutes and delivered during business hours at a private outpatient α-rTMS centre in North Brisbane, Australia.

#### Level 2 polysomnography (L2PSG)

This study adapted the PSG device set-up as previously described by Russo et al. (2021). Briefly, the PSG was performed using a Nox-A1 device (Nox Medical, Iceland). Monitored signals included frontal electroencephalogram (EEG: FpZ, F4, and F3), two electrooculograms (EOG), two chin electromyogram (EMG), electrocardiogram, chest respiratory inductance plethysmography (cRIP), pulse oximetry (Nonin3150 with one of 2 age-appropriately sized probes) and audio. Airflow was assessed using cRIP flow, a calculated signal derived from the respiratory bands.

The pulse oximetry connects to the main device via Bluetooth. All the leads were connected to a battery-powered device worn on the child’s chest and were bundled together to prevent a choking hazard. The pulse oximetry was worn on the less dominant wrist using medical tape, and the probe was secured on the middle finger using an adhesive, non-woven fabric. Data was scored by a paediatric sleep scientist (JG) according to the guidelines of the American Academy of Sleep Medicine (Berry, 2014) and reviewed by a paediatric sleep physician (SB).

The signal quality of this study used similar criteria as Russo et al. (2021). The Noxturnal software-rated signal quality was assessed and confirmed manually by a sleep specialist (SB). The RIP bands were rated as inadequate if either the respiratory band signal was absent or a poor signal for >10% of sleep time. SB manually assessed the EEG, EOG, and chin EMG for quality. To be acceptable, all 3 EEG signals had to be present and sufficiently free from any artefact for ≥ 90% of sleep such that sleep stages could be evaluated. The EOG was assessed as adequate if an artefact-free signal was present for > 90% of sleep time and REMs could be identified. Chin EMG was assessed as adequate if present for ≥ 90% of the recording, and there was appropriate differentiation between wake, REM

#### sleep and NREM sleep

L2PSG was administered at the different study phase. The setup was conducted in the participant’s home and at an average of 60 minutes before their usual bedtime. The device was set to start automatically at the child’s routine bedtime. Also, a Nox-A1 app on an iPad was provided for primary caregivers to intermittently visualise and adjust any leads with poor connectivity and impedance levels overnight. The primary caregivers were provided with an on-call mobile number to contact a trained sleep technician (UE) anytime. Primary caregivers were interviewed about when the child fell asleep and woke up and any behaviours around compliance that were observed overnight.

### Assessment of study adequacy

The L2PSG data were deemed as adequate when assessed for > 6h of sleep, and key channels (EEG, RIP bands, cRIP airflow and pulse oximetry) present for at least 90% of sleep.

### Costs

The cost of a one-night L2PSG was determined to be $AUD 258 and detailed as follows: $AUD 23 for RIP belt, $AUD 8 for consumables (such as alcohol prep wipes, 25mm and 30mm electrode dots, and lithium batteries), ∼ $AUD 23 as travel cost of PhD student to each participant’s home (0.55c per km) and $AUD 54 per hour for set up, and $AUD 150 per analysis of recording. The cost of a Nox A1 was $AUD 12,500, but this was provided by SB.

#### Statistical analysis

Descriptive statistics for clinical characteristics and sleep parameters are expressed as mean, and standard deviation, and interview responses are displayed as frequencies and percentages.

## Result

Table 1 summarises participant demographic data at baseline.

**Table 1:** Participant’s demographic characteristics.

| Baseline characteristics | TG (n = 10) | WLCG (n = 10) |
| --- | --- | --- |
| *Demographics* |  |  |
| Mean age (SD)/range | 9 (1.49)/ 7–12 | 9.2 (1.68)/ 6–11 |
| Boys (n) | 7 (70%) | 9 (90%) |
| Girls (n) | 3 (30%) | 1 (10%) |
| Years since ASD diagnosis (SD)/range | 2.9 (1.79)/1–6 | 3 (1.82)/1–6 |
| *Primary caregiver* |  |  |
| Mother | 10 | 9 |
| Grandmother | 0 | 1 |
| *Reported sleep difficulties* |  |  |
| Bedtime resistance | 7 | 6 |
| Sleep anxiety | 5 | 3 |
| Prolonged sleep onset latency | 7 | 4 |
| Nighttime waking | 4 | 4 |
| Early waking/short sleep | 6 | 5 |
| Daytime tiredness/sleepiness | 3 | 3 |
| *Other comorbidities* |  |  |
| Attention Deficit Hyperactive Disorder | 7 | 7 |
| Anxiety (General anxiety disorder, social anxiety) | 6 | 4 |
| Dyslexia/ Dyscalculia/ Dysgraphia/sensory disorder | 3 | 2 |
| *Existing medications* |  |  |
| Yes | 8 | 6 |
| No | 2 | 4 |
| α - 2a adrenergic agonist (clonidine, guanfacine) | 5 | 2 |
| Melatonin | 4 | 3 |
| Stimulant (Lis dexamphetamine, methylphenidate) | 6 | 5 |
| *Existing allied health therapies* |  |  |
| Counsellor/psychotherapy/psychology | 8 | 4 |
| Exercise physiology/physiotherapy | 4 | 1 |
| Occupational therapy | 10 | 5 |
| Speech therapy | 4 | 3 |

A total of 20 children (mean age ±SD = 9.1 ±1.55), comprising 16 boys and 4 girls diagnosed with autism spectrum disorder (level 2), were recruited for this study. Participants had an average of 3 years following ASD diagnosis. The most frequently reported sleep difficulties by primary caregivers were bedtime resistance (n=13, 65%), prolonged sleep onset latency (n=11, 55%), early waking/short sleep (n=11, 55%), sleep anxiety (n=8, 40%), nighttime waking (n=8, 40%), and daytime tiredness/sleepiness (n=6, 30%),.

Reported co-diagnoses were attention deficit hyperactivity disorder (n=14, 70%), anxiety (n=10, 50%), and sensory or learning difficulties (n=5, 25%). Most participants were on medication (n=14, 70%) such as stimulants (n=11, 55%), α - 2a adrenergic agonist (n=7, 35%), and melatonin (n=7, 35%), while frequently accessed allied therapies were occupational (n=15, 75%), counsellor/psychotherapy/psychology (n= 12, 60%), speech (n=7, 35%), and exercise physiology/physiotherapy (n=5, 25%).

Table 2 shows that of the proposed 90 L2PSG, 80 were feasible. The other 10 L2PSG were infeasible due to drop-out or missed studies arising from sickness (n=1, 5% at T4), family relocation (n=1, 5% at T5), change in family dynamics (n=1, 5% at T5) and withdrawal to enable further medical investigation unrelated to the study (n=2, at T3 (20%), T4 (10%) and T5 (10%)). One participant had a skin allergy to the 25mm electrode dots used in the device setup and dropped out at T5.

**Table 2:** Feasible and adequate home (level 2) PSG at each time point.

| Participants # | Groups | Baseline<br>(T1)<br>(n = 20) | Post-intervention<br>(T2)<br>(n = 20) | Post-intervention<br>(T3)<br>(n = 8) | One-month post-<br>study follow-up<br>(T4)<br>(n = 17) | Four-month post-<br>study follow-up<br>(T5)<br>(n = 15) |
| --- | --- | --- | --- | --- | --- | --- |
| 1 | Treatment<br>group | ++ | ++ |  | ++ | ++ |
| 2 |  | ++ | ++ |  | ++ | ++ |
| 3 |  | ++ | ++ |  | ++ | ++ |
| 4 |  | ++ | ++ |  | ++ | ++ |
| 5 |  | ++ | ++ |  | ++ | ++ |
| 6 |  | ++ | ++ |  | ++ | ++ |
| 7 |  | ++ | ++ |  | ++ | ++ |
| 8 |  | ++ | ++ |  | +- | xx |
| 9 |  | ++ | +- |  | +- | ++ |
| 10 |  | +- | ++ |  | ++ | +- |
| 11 | Waitlist<br>control<br>group | ++ | ++ | ++ | ++ | ++ |
| 12 |  | ++ | ++ | ++ | ++ | ++ |
| 13 |  | ++ | ++ | ++ | ++ | ++ |
| 14 |  | ++ | ++ | ++ | ++ | ++ |
| 15 |  | ++ | ++ | ++ | +- | ++ |
| 16 |  | ++ | ++ | ++ | ++ | xx |
| 17 |  | ++ | ++ | ++ | xx | ++ |
| 18 |  | ++ | ++ | ++ | ++ | xx |
| 19 |  | ++ | ++ | xx | xx | xx |
| 20 |  | ++ | ++ | xx | xx | xx |

Key: Feasible and adequate (++); feasible but inadequate (+-); missed or dropped out (xx)

Of the 80 administered L2PSG studies, 74 (93%) were deemed technically adequate (Table 3). Time 1 and 2 (T1 and T2) were 95% (n=19/20) while T3, T4 and T5 were 100% (n=8/8), 82% (n=14/17), and 93% (n=14/15) adequate, respectively (Table 3). The six (7%) inadequate L2PSGs were all due to >90% EEG absence/artefact. The multiple L2PSG showed that participants did not have a sleep disorder requiring medical attention, except for one participant at time T3, who had mild obstructive sleep apnoea. With 80 L2PSG administered, the total cost is estimated at $AUD 20,640.

**Table 3:**
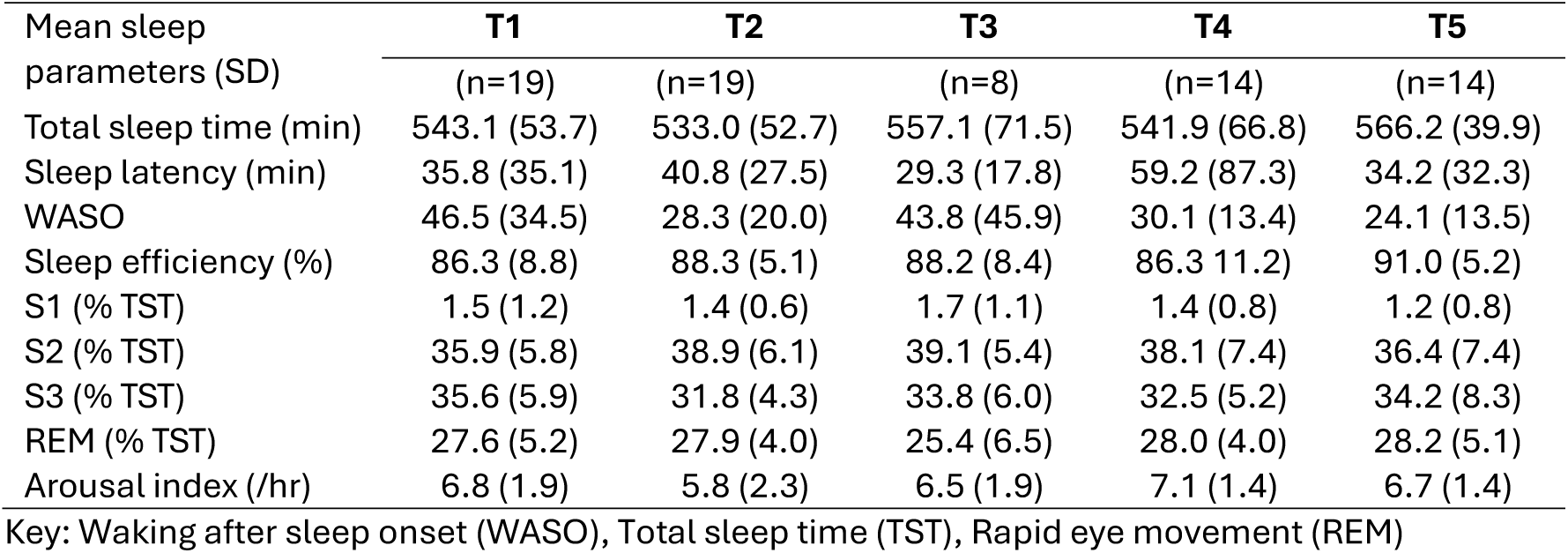
Mean and standard deviation of sleep parameters at each study time point.

### Primary caregiver’s report

At Time 1, primary caregivers reported that two (10%) participants had experienced discomfort with the setup overnight, but no significant difficulties or events. All primary caregivers found using the Nox-A1 app on an iPad intuitive and useful for adjusting signal quality (only two calls were to on-call support during the research with questions regarding EEG signals), and the L2PSG setup was relatively easy and convenient for them and their children.

## Discussion

This prospective observational study demonstrates the feasibility and adequacy of Level 2 PSG (L2PSG) during an interventional research involving children with autism spectrum disorder (ASD) and sleep difficulties.

The majority of L2PSG administered in this study were feasible. In specific, the first L2PSG at T1 was 100% feasible despite being administered to participants without a desensitisation technique (Kawai et al., 2022) or an adaptation L2PSG (Jiao et al., 2022). Other non-interventional L2PSG studies involving children with ASD reported a feasibility of 81-91% (Ioan et al., 2020, Russo et al., 2021, Marcus et al., 2014). The high feasibility in this study may collectively involve setting up the PSG at home, an hour before the participant’s routine bedtime and indirect airflow monitoring, i.e., chest RIP. Most primary caregivers reported that the set-up at home was relatively easy and convenient for them and their children. Setting up closer (∼1hr) to bedtime than in the early afternoon at a clinic may have minimised disruption to the child’s typical bedtime routine and reduced any potential for anxiety and overstimulation following transportation to and from the clinic (Henderson et al., 2011). More so, monitoring airflow via cRIP than the poorly tolerated nasal prongs and oronasal thermistor (Lanzlinger et al., 2023) may have prevented the heightening of tactile sensitivity, a significant driver of sleep difficulties within the population (Tzischinsky et al., 2018). However, it must be noted that the maintenance of a 95-100% feasibility following repeated L2PSG (T2-T5) may be due to the participants becoming more tolerant of the set-up and or the effect of the intervention on tactile sensitivity. In an open-label study involving 39 children, Gao et al. (2022) showed that the intervention, repetitive transcranial magnetic stimulation, improved sensory abnormalities, especially tactile sensitivity. Although there were reported discomforts experienced by a few participants, this did not lead to early termination or a decline in repeated L2PSG, except for one participant who declined future L2PSG due to a skin allergy to the sticky dots. Such high feasibility of L2PSG, especially over repeated timepoints, is significant in interventional research as a low feasibility due to L2PSG related dropout lead to poor generalisability of the result and study quality (Leon et al., 2006).

Further, the signal quality from most participants in this study was adequate for critical sleep parameters such as total sleep time (TST), sleep latency, waking after sleep onset (WASO), sleep efficiency (SE), arousal index and sleep stagings. The signal quality was also adequate to capture mild obstructive sleep apnea (OSA) in one participant, supporting the findings from the L2PSG study by Russo et al. (2021) in which cRIP was used in diagnosing OSA in children with or without neurodevelopmental conditions. Although there were a few inadequate L2PSG, all due to EEG artefact/absence. This is not uncommon and has been reported in other L2PSG (Russo et al., 2021) and L1PSG studies (Lanzlinger et al., 2023). The high adequacy of this study may have been supported by primary caregivers addressing any poor signal quality in real-time using the Nox-A1 app provided on an iPad, thereby ensuring high-quality signal acquisition at par with L1PSG (Portier et al., 2000)

Therefore, it could be suggested that this study method is optimal for signal acquisition during research requiring an objective assessment of sleep. To illustrate this, primary caregivers frequently reported sleep difficulties such as bedtime resistance, sleep anxiety, prolonged sleep onset latency, nighttime waking, and early waking, among others (Rana et al., 2021, Maurer et al., 2023), however, the L2PSG studies showed none of the children had a sleep disorder requiring medical attention. To support this, all sleep parameters in this study, except for the low S1%, were within the range of typically developing children of similar age who underwent non-interventional L1PSG (Scholle et al., 2011). A low S1% may suggest the child transitioned faster to deep sleep, which may support reports by primary caregivers of some or most children sleeping well or better than usual during non-interventional L2PSG (Goodwin et al., 2001, Russo et al., 2021).

A meta-analysis concluded that different sleep parameters were affected by different factors (Chen et al., 2021). However, in a non-interventional L2PSG, autistic children showed a lower S1% in addition to lower TST, higher S3%, lower REM% and higher REM latency compared to typically developing children of similar age (Kawai et al., 2022). The additional difference in sleep parameters compared to this study may be due to the heterogeneity in participants’ demography, such as age and gender or comorbidities, such as attention deficit hyperactivity disorder (ADHD). For instance, compared to this study, Kawai et al. (2022) recruited autistic children between the ages of 4 and 18 years, with a lower female-to-male ratio and fewer ADHD comorbidities. Such a wide age range may impact the TST, given that this differs across age ranges (Banaschewski et al., 2022).

Alternatively, the longer TST in this study may correlate with the high number of females with ADHD. A recent non-interventional L1PSG study concluded on a potential sex-specific difference impacting sleep parameters, as females with ADHD showed a significantly longer TST than males (Lin et al., 2025). In another L2PSG, the low female-to-male ratio of children with ADHD showed lower S1% in addition to lower TST, higher S3%, and lower REM% compared to typically developing controls (Gruber et al., 2009). Given the prevalence of ASD and comorbid ADHD in PSG studies (Kawai et al., 2022, Russo et al., 2021, Kevat et al., 2025, Lanzlinger et al., 2023) and the overlap between sleep difficulties in both conditions, it may be challenging for researchers to distinguish clearly and research separately the relevant differences between both conditions (Chen et al., 2021).

Put together, the high feasibility and adequacy support the financial benefit of L2PSG compared to L1PSG for research purposes (Portier et al., 2000). It is recommended that the cost difference between L1PSG and L2PSG be considered in the context of a 20% failure in the latter (Portier et al., 2000). This interventional study had a 7% failure rate across 80 L2PSGs, and the cost of one L2PSG ($AUD 258) was relatively low compared to previous non-interventional PSG studies. For instance, L2PSG costs $AUD 814.35 (Carpentier et al., 2014) while another study reported $AUD 354.82 (Portier et al., 2000). L1PSG was estimated to cost $AUD 974.45 (Pelletier-Fleury et al., 1998) or $AUD 1,196.39 (Carpentier et al., 2014) (All c*urrencies were in Euros, corrected for inflation and converted to Australian dollars using 2025 rates)*.

This study method of training primary caregivers to address poor signal quality in real time may have minimised failure rates, thereby saving the cost per hour in employing a trained staff during L1PSG. Withers et al. (2022) showed that L2PSG needed a total of 2.6 hours of staff time as compared to L1PSG, which required 8.6 hours. Using the staff cost of this study, an L1PSG is estimated to cost $AUD 685.3 ($AUD 77 x 8.6hrs) in staff time. The additional cost of travelling to the participant’s home to set up and returning the next day to pick up the device may have had a positive impact on the retention rate in the study. With multiple L2PSGs at different study timepoints, if the cost of transportation were on participants, this may cause some to drop out, especially due to the high cost of living or other life commitments. Further, this study was cost-effective in achieving a quality RCT given that the infeasible and inadequate L2PSG were less than 17%, which is comparable to other high quality RCT (Wood et al., 2004)

### Study strengths and limitations

This is the first study to observe the feasibility and adequacy of level 2 polysomnography (L2PSG) in an interventional randomised controlled trial involving children with autism spectrum disorder and sleep difficulties. Only a single L2PSG (1%) was infeasible due to the participant not tolerating the setup. The signal quality was highly adequate despite the sensory sensitivity reported in the population. A cost analysis for 80 L2PSG ($AUD 20,640) was estimated. This study shows that L2PSG may be a feasible and adequate approach to sleep studies, especially in interventional research with concerns of heightened sensory sensitivities and intolerance within the population (Gao et al., 2021). More so, the considerably low cost of L2PSG may enable interventional research requiring repeated measurements or long-term follow-ups. The 4:1 ratio of boys to girls parallels those reported in epidemiological studies (Elsabbagh et al., 2012, Maenner et al., 2023) and may support the external validity of this study.

However, the small sample size and recruitment of participants from a specific age (6-12 yrs) and ASD diagnosis (level 2) may limit its external validity due to the heterogeneous nature of the ASD population. For instance, children diagnosed with ASD level 3 may be more sensitive and less tolerant of L2PSG. Other factors, such as how comorbid ADHD, medications and other allied health therapies contribute to the feasibility and adequacy of L2PSG, are not fully understood. Further, anthropometric measures such as height, weight, and neck circumference were not collected, which would have enabled correlation with the diagnosis of obstructive sleep apnoea within the population (Barış et al., 2017). This study is also limited by the lack of periodic limb movement during sleep (PLMS) measurement and synchronised video monitoring for observing parasomnia and potentially external disturbances like noise or family interaction that may impact the scoring quality. Comparing L2PSG and L1PSG during an interventional study may potentially further the debate.

### Summary

In summary, evidence from this study adds to the increasing interest in L2PSG in children with ASD (Russo et al., 2021, Kawai et al., 2022) and its potential use in interventional research. However, the feasibility and adequacy of L2PSG during an interventional study involving children with ASD may be predicated on the complexity of the child’s sleep difficulties, safety and family dynamics in the child’s home, participant’s allergies, high-quality equipment, primary caregiver’s ability to address signal issues in real-time, and availability of on-call supports. The financial cost of L2PSG will depend on the sample size, relative distance between participants’ homes (if multiple setups are to be done per night), number of trained researchers for setup and the cost of transportation between participants’ homes.

### Recommendation

Given feasibility, adequacy and cost-effectiveness of L2PSG in this interventional research involving children with ASD, multiple studies on a larger sample size and using the described methodol are required. In addition, a set-up coupled with synchronised video monitoring may optimise the the scoring quality.

### Conclusion

The study indicates the feasibility, adequacy and cost-effectiveness of level 2 polysomnography involving children with ASD during an interventional randomised controlled trial. This preliminary study provides valuable insights into the field of paediatric sleep medicine. Repeats of this method using diverse ASD population and age and larger sample sizes are warranted.

## Data Availability

All data produced in the present study are available upon reasonable request to the authors.

## Reference

Aathira, R., Gulati, S., Tripathi, M., Shukla, G., Chakrabarty, B., Sapra, S., Dang, N., Gupta, A., Kabra, M. & Pandey, R. M. 2017. Prevalence of sleep abnormalities in Indian children with autism spectrum disorder: a cross-sectional study. Pediatric Neurology, 74, 62–67.

Banaschewski, T., Bruni, O., Fuentes, J., Hill, C. M., Hvolby, A., Posserud, M.-B. & Schroder, C. 2022. Practice tools for screening and monitoring insomnia in children and adolescents with autism spectrum disorder. Journal of Autism and Developmental Disorders, 52, 3758–3768.

Barış, H. E., Gökdemir, Y., Eralp, E. E., İkizoğlu, N. B., Karakoç, F., Karadağ, B. & Ersu, R. 2017. Clinical and polysomnographic features of children evaluated with polysomnography in pediatric sleep laboratory. Turk Pediatri Ars, 52, 23–29.

Berry, R. B. 2014. The AASM manual for the scoring of sleep and associated events: rules, terminology and technical specifications. version 2.1. Darien Illinois: American Academy of Sleep Medicine.

Bessey, M., Richards, J. & Corkum, P. 2013. Sleep Lab Adaptation in Children with Attention-Deficit/Hyperactivity Disorder and Typically Developing Children. Sleep Disorders, 2013, 698957.

Braun, M., Stockhoff, M., Tijssen, M., Dietz-Terjung, S., Coughlin, S. & Schöbel, C. 2024. A systematic review on the technical feasibility of home-polysomnography for diagnosis of sleep disorders in adults. Current Sleep Medicine Reports, 10, 276–288.

Brockmann, P. E., Perez, J. L. & Moya, A. 2013. Feasibility of unattended home polysomnography in children with sleep-disordered breathing. International journal of pediatric otorhinolaryngology, 77, 1960–1964.

Buckley, A., Wingert, K., Swedo, S., Thurm, A., Sato, S., Appel, S. & Rodriguez, A. J. 2013. First night effect analysis in a cohort of young children with autism spectrum disorder. Journal of Clinical Sleep Medicine, 9, 67–70.

Campbell, A. J. & Neill, A. M. 2011. Home set-up polysomnography in the assessment of suspected obstructive sleep apnea. Journal of sleep research, 20, 207–213.

Carpentier, N., Jonas, J., Schaff, J.-L., Koessler, L., Maillard, L. & Vespignani, H. 2014. The feasibility of home polysomnographic recordings prescribed for sleep-related neurological disorders: a prospective observational study. Neurophysiologie Clinique/Clinical Neurophysiology, 44, 251–255.

Chen, X., Liu, H., Wu, Y., Xuan, K., Zhao, T. & Sun, Y. 2021. Characteristics of sleep architecture in autism spectrum disorders: A meta-analysis based on polysomnographic research. Psychiatry Research, 296, 113677.

Douglas, J. A., Chai-Coetzer, C. L., McEvoy, D., Naughton, M. T., Neill, A. M., Rochford, P., Wheatley, J. & Worsnop, C. 2017. Guidelines for sleep studies in adults-a position statement of the Australasian Sleep Association. Sleep Med, 36, S2–S22.

Elsabbagh, M., Divan, G., Koh, Y. J., Kim, Y. S., Kauchali, S., Marcín, C., Montiel-Nava, C., Patel, V., Paula, C. S. & Wang, C. 2012. Global prevalence of autism and other pervasive developmental disorders. Autism research, 5, 160–179.

Ezedinma, U., Swierkowski, P. & Fjaagesund, S. 2022. Outcomes from Individual Alpha Frequency Guided Repetitive Transcranial Magnetic Stimulation in Children with Autism Spectrum Disorder–A Retrospective Chart Review. Child Psychiatry & Human Development, 1-10.

Feng, S., Huang, H., Wang, N., Wei, Y., Liu, Y. & Qin, D. 2021. Sleep disorders in children with autism spectrum disorder: Insights from animal models, especially non-human primate model. Frontiers in Behavioral Neuroscience, 15, 673372.

Gao, L., Wang, C., Song, X. R., Tian, L., Qu, Z. Y., Han, Y. & Zhang, X. 2021. The Sensory Abnormality Mediated Partially the Efficacy of Repetitive Transcranial Magnetic Stimulation on Treating Comorbid Sleep Disorder in Autism Spectrum Disorder Children. Front Psychiatry, 12, 820598.

Gao, L., Wang, C., Song, X. R., Tian, L., Qu, Z. Y., Han, Y. & Zhang, X. 2022. The Sensory Abnormality Mediated Partially the Efficacy of Repetitive Transcranial Magnetic Stimulation on Treating Comorbid Sleep Disorder in Autism Spectrum Disorder Children. Frontiers in Psychiatry, 12.

Ghegan, M. D., Angelos, P. C., Stonebraker, A. C. & Gillespie, M. B. 2006. Laboratory versus portable sleep studies: a meta-analysis. The Laryngoscope, 116, 859–864.

Goodwin, J. L., Enright, P. L., Kaemingk, K. L., Rosen, G. M., Morgan, W. J., Fregosi, R. F. & Quan, S. F. 2001. Feasibility of using unattended polysomnography in children for research— report of the Tucson Children’s Assessment of Sleep Apnea study (TuCASA). Sleep, 24, 937–944.

Gruber, R., Xi, T., Frenette, S., Robert, M., Vannasinh, P. & Carrier, J. 2009. Sleep disturbances in prepubertal children with attention deficit hyperactivity disorder: a home polysomnography study. Sleep, 32, 343–350.

Henderson, J. A., Barry, T. D., Bader, S. H. & Jordan, S. S. 2011. The relation among sleep, routines, and externalizing behavior in children with an autism spectrum disorder. Research in Autism Spectrum Disorders, 5, 758–767.

Hodge, D., Parnell, A. M., Hoffman, C. D. & Sweeney, D. P. 2012. Methods for assessing sleep in children with autism spectrum disorders: A review. Research in Autism Spectrum Disorders, 6, 1337–1344.

Ioan, I., Weick, D., Schweitzer, C., Guyon, A., Coutier, L. & Franco, P. 2020. Feasibility of parent-attended ambulatory polysomnography in children with suspected obstructive sleep apnea. Journal of Clinical Sleep Medicine, 16, 1013–1019.

Jiao, J., Tan, L., Zhang, Y., Li, T. & Tang, X. 2022. Repetitive transcranial magnetic stimulation for insomnia in patients with autism spectrum disorder: Study protocol for a randomized, double-blind, and sham-controlled clinical trial. Frontiers in Psychiatry, 13, 977341.

Kawai, M., Buck, C., Chick, C. F., Anker, L., Talbot, L., Schneider, L., Linkovski, O., Cotto, I., Parker-Fong, K. & Phillips, J. 2022. Sleep architecture is associated with core symptom severity in autism spectrum disorder. Sleep.

Kevat, A., Alwadhi, D., Collaro, A., Bernard, A., Vandeleur, M., Waters, K. & Chawla, J. 2025. Parent-reported experiences of in-laboratory polysomnography in children with neurodevelopmental disorders: A cross-sectional multi-centre study. Sleep and Breathing, 29, 1–8.

Lanzlinger, D., Kevat, A., Collaro, A., Poh, S. H., Pérez, W. P. & Chawla, J. 2023. Tolerance of polysomnography in children with neurodevelopmental disorders compared to neurotypical peers. Journal of Clinical Sleep Medicine, 19, 1625–1631.

Leon, A. C., Mallinckrodt, C. H., Chuang-Stein, C., Archibald, D. G., Archer, G. E. & Chartier, K. 2006. Attrition in randomized controlled clinical trials: methodological issues in psychopharmacology. Biological psychiatry, 59, 1001–1005.

Lin, C.-H., Wu, P.-Y., Hong, S.-Y., Chang, Y.-T., Lin, S.-S. & Chou, I.-C. 2025. The Role of Polysomnography for Children with Attention-Deficit/Hyperactivity Disorder. Life, 15, 678.

Maenner, M. J., Warren, Z., Williams, A. R., Amoakohene, E., Bakian, A. V., Bilder, D. A., Durkin, M. S., Fitzgerald, R. T., Furnier, S. M., Hughes, M. M., Ladd-Acosta, C. M., McArthur, D., Pas, E. T., Salinas, A., Vehorn, A., Williams, S., Esler, A., Grzybowski, A., Hall-Lande, J., Nguyen, R. H. N., Pierce, K., Zahorodny, W., Hudson, A., Hallas, L., Mancilla, K. C., Patrick, M., Shenouda, J., Sidwell, K., DiRienzo, M., Gutierrez, J., Spivey, M. H., Lopez, M., Pettygrove, S., Schwenk, Y. D., Washington, A. & Shaw, K. A. 2023. Prevalence and Characteristics of Autism Spectrum Disorder Among Children Aged 8 Years – Autism and Developmental Disabilities Monitoring Network, 11 Sites, United States, 2020. MMWR Surveill Summ, 72, 1–14.

Marcus, C. L., Traylor, J., Biggs, S. N., Roberts, R. S., Nixon, G. M., Narang, I., Bhattacharjee, R., Davey, M. J., Horne, R. S. & Cheshire, M. 2014. Feasibility of comprehensive, unattended ambulatory polysomnography in school-aged children. Journal of Clinical Sleep Medicine, 10, 913–918.

Maski, K., Holbrook, H., Manoach, D., Hanson, E., Kapur, K. & Stickgold, R. 2015. Sleep dependent memory consolidation in children with autism spectrum disorder. Sleep, 38, 1955–1963.

Maurer, J. J., Choi, A., An, I., Sathi, N. & Chung, S. 2023. Sleep disturbances in autism spectrum disorder: Animal models, neural mechanisms, and therapeutics. Neurobiology of Sleep and Circadian Rhythms, 14, 100095.

Miano, S., Bruni, O., Elia, M., Trovato, A., Smerieri, A., Verrillo, E., Roccella, M., Terzano, M. G. & Ferri, R. 2007. Sleep in children with autistic spectrum disorder: a questionnaire and polysomnographic study. Sleep medicine, 9, 64–70.

Mindell, J. A., Meltzer, L. J., Carskadon, M. A. & Chervin, R. D. 2009. Developmental aspects of sleep hygiene: findings from the 2004 National Sleep Foundation Sleep in America Poll. Sleep medicine, 10, 771–779.

Moore, M., Evans, V., Hanvey, G. & Johnson, C. 2017. Assessment of sleep in children with autism spectrum disorder. Children, 4, 72.

Newell, J., Mairesse, O., Verbanck, P. & Neu, D. 2012. Is a one-night stay in the lab really enough to conclude? First-night effect and night-to-night variability in polysomnographic recordings among different clinical population samples. Psychiatry research, 200, 795–801.

O’Sullivan, R., Bissell, S., Hamilton, A., Bagshaw, A. & Richards, C. 2023. Concordance of objective and subjective measures of sleep in children with neurodevelopmental conditions: A systematic review and meta-analysis. Sleep Medicine Reviews, 101814.

Pedersen, M. J., Leonthin, H., Mahler, B., Rittig, S., Jennum, P. J. & Kamperis, K. 2023. Two nights of home polysomnography in healthy 7-14-year-old children–Feasibility and intraindividual variability. Sleep Medicine, 101, 87–92.

Pelletier-Fleury, N., Fontanille, C., Lanoe, J. & Fleury, B. 1998. Mesure des coûts réels de la polysomnographie au laboratoire de sommeil. Rev Mal Respir, 15, 1S75.

Portier, F., Portmann, A., Czernichow, P., Vascaut, L., Devin, E., Benhamou, D., Cuvelier, A. & Muir, J. F. 2000. Evaluation of home versus laboratory polysomnography in the diagnosis of sleep apnea syndrome. American journal of respiratory and critical care medicine, 162, 814–818.

Rana, M., Kothare, S. & DeBassio, W. 2021. The Assessment and Treatment of Sleep Abnormalities in Children and Adolescents with Autism Spectrum Disorder: A Review. J Can Acad Child Adolesc Psychiatry, 30, 25–35.

Russo, K., Greenhill, J. & Burgess, S. 2021. Home (Level 2) polysomnography is feasible in children with suspected sleep disorders. Sleep Medicine, 88, 157–161.

Scholle, S., Beyer, U., Bernhard, M., Eichholz, S., Erler, T., Graneß, P., Goldmann-Schnalke, B., Heisch, K., Kirchhoff, F. & Klementz, K. 2011. Normative values of polysomnographic parameters in childhood and adolescence: quantitative sleep parameters. Sleep medicine, 12, 542–549.

Tzischinsky, O., Meiri, G., Manelis, L., Bar-Sinai, A., Flusser, H., Michaelovski, A., Zivan, O., Ilan, M., Faroy, M. & Menashe, I. 2018. Sleep disturbances are associated with specific sensory sensitivities in children with autism. Molecular autism, 9, 1–10.

Withers, A., Maul, J., Rosenheim, E., O’Donnell, A., Wilson, A. & Stick, S. 2022. Comparison of home ambulatory type 2 polysomnography with a portable monitoring device and in-laboratory type 1 polysomnography for the diagnosis of obstructive sleep apnea in children. Journal of Clinical Sleep Medicine, 18, 393–402.

Wood, A. M., White, I. R. & Thompson, S. G. 2004. Are missing outcome data adequately handled? A review of published randomized controlled trials in major medical journals. Clinical trials, 1, 368–376.

